# Health-Seeking Behavior and Anxiety of Cancer Patients in Bangladesh during the COVID-19 Pandemic: A Cross-Sectional Study

**DOI:** 10.1101/2024.08.19.24312282

**Authors:** Nur-A-Safrina Rahman, Munmun Mustafa, Tahsin Tasneem Tabassum, Sumona Haque Simu, Mridul Gupta, Sumaiya Afrin, Maisha Samiha, Shahra Tanjim Moulee, Faisal Abdullah, Sifat Sharmin, Bilkis Akhter Loken, Sadia Mahmud Trisha, Md. Saimon, Vivek Podder, Priya Singhania, ANM Shamsul Islam

## Abstract

**Background:** The COVID-19 pandemic has posed unique challenges for cancer patients, who not only require ongoing medical care but also face an elevated risk of infection. Investigating the health-seeking behavior and barriers among adult cancer patients during this global crisis is crucial for ensuring their access to essential care amidst the pandemic’s complexities.

**Objective:** This cross-sectional study aimed to assess the health-seeking behavior, perceived barriers, and anxiety among adult cancer patients during the COVID-19 pandemic.

**Materials and Methods:** The study, conducted from August 2020 to December 2020, involved 210 participants purposively selected from the National Institute of Cancer Research and Hospital and Ahsania Mission Cancer and General Hospital in Dhaka. Data was collected through face-to-face interviews using a pre-tested semi-structured questionnaire and analyzed using SPSS (Version 26).

**Results:** Among the 210 participants, 56.2% were male, 28.6% aged 46-55, and 36.7% had no formal education. Approximately 52.3% preferred public healthcare facilities, while 6.2% sought homeopathy or pharmacy advice for symptoms. Significant differences emerged in post-pandemic healthcare provider contact (p-0.0). Notably, 88.1% missed appointments, with 78.3% taking no action. Barriers included transport issues (77.1%), reduced income (59%), and lacking financial (53.4%) and mental support (56.6%). Conversely, respondents downplayed public awareness (80%), infection risk from others (84.7%), healthcare provider infection risk (82.4%), and hospital overcrowding (64.8%). Fear of hospitals correlated with public awareness (p-0.0). On the GAD-7 scale, most had minimal anxiety (53.8%), with a weak provider contact-anxiety correlation (p-0.03). Healthcare providers excelled in precautions (99.5%) and health status communication (85.3%).

**Conclusion:** Despite the barriers and risks posed by the pandemic, cancer patients prioritized their care. Given the need for continued cancer care and the elevated risk of COVID-19 among cancer patients, adapting measures to align with the population’s real needs could prove highly beneficial.

## Introduction

In January 2020, the World Health Organization (WHO) declared a global public health emergency in response to the rapid emergence of the COVID-19 pandemic [1]. Within six months, this novel virus had left an indelible mark, with approximately 20 million confirmed cases and a staggering 700,000 deaths reported worldwide [1]. This unfolding health crisis brought forth a myriad of concerns and responses, profoundly altering health-seeking behaviors among individuals globally.

In the wake of COVID-19, concerns about the risk of viral exposure, limited access to traditional healthcare services, and the deluge of often contradictory information led to a notable shift in health-seeking behavior. As individuals found themselves confined to their homes and starved of reliable, in-person medical advice, the Internet emerged as their primary source of information. However, amid this turbulent landscape, a particularly vulnerable group emerged -individuals already grappling with the formidable challenge of cancer. The COVID-19 pandemic posed a heightened threat to these patients, not only due to their immunocompromised state resulting from malignancy and anti-cancer therapies such as chemotherapy and surgery [2], but also due to the startling statistic that approximately 5.6% of cancer patients succumbed to the virus, compared to a 2.6% fatality rate among non-cancer patients [3]. Cancer patients, by nature, straddle the realm of both non-communicable and communicable diseases. During this unprecedented period, the urgency of continuous treatment and specialized care for cancer patients was magnified manifold due to their heightened vulnerability.

Within the context of Bangladesh, a country grappling with the complex interplay of a burgeoning population, economic constraints, and healthcare challenges, the collision of the COVID-19 pandemic with the pre-existing cancer burden has had profound implications. With a population of 142 million, Bangladesh ranks as the ninth most populous nation globally, classified as a low-middle-income country [4]. Here, an estimated 13 to 15 lakh cancer patients navigate their healthcare journey, with an additional 2 lakh new cancer diagnoses annually [5]. However, the arrival of the COVID-19 pandemic prompted a paradigm shift in healthcare dynamics within the nation.

COVID-19 has exposed the challenges healthcare systems face in low—and middle-income countries (LMICs) in delivering efficient services. The pandemic has exacerbated the pre-existing strain on primary healthcare systems, siphoning resources toward the urgent pandemic response at the expense of routine healthcare services. This diversion has inevitably impeded access to healthcare, a phenomenon that has not gone unnoticed among the populace.

Healthcare-seeking behaviour, often characterised by the actions taken by individuals in response to perceived illness or disease states, is a critical determinant of health outcomes. It encompasses a broad spectrum of behaviours, including the time elapsed between symptom onset and seeking medical care, the choice of healthcare providers, adherence to prescribed treatments, and the myriad of reasons influencing these decisions [6]. Of concern is the impact of health-seeking behavior on cancer outcomes, where early diagnosis can be the difference between successful treatment and disease progression. Typically, the diagnosis of cancer occurs when individuals manifest symptoms and subsequently seek medical evaluation. However, the early stages of the COVID-19 pandemic saw a deviation from this norm, as health-seeking behaviors among the population were heavily influenced by government directives. For cancer patients already navigating a complex and often daunting healthcare journey, the confluence of factors such as lockdowns, transportation constraints, and the fear of nosocomial COVID-19 transmission posed significant hurdles. Regrettably, these hurdles resulted in delayed or missed appointments, and in some instances, patients opted to forgo essential medical care. As cancer patients, they stood not only to lose access to vital treatments but also to face the unintended consequence of disease progression. Furthermore, the pandemic introduced an additional layer of complexity to the psychological well-being of cancer patients. The alteration of cancer care routines, treatment delays, and the cancellation of non-urgent procedures fueled anxiety and fear among this cohort, a phenomenon exacerbated by heightened health anxiety. These individuals often misinterpreted benign bodily sensations as signs of grave illness, leading to increased stress, compromised decision-making, and an array of adverse behavioral responses.

Recognising the critical need for cancer patients to maintain uninterrupted care, including prevention measures and routine alterations necessitated by the pandemic, the COVID-19 crisis presents a unique opportunity. It affords an unprecedented lens through which to study the health-seeking behaviors faced by adult cancer patients during this unparalleled moment in medical history. Therefore, we aim to investigate the health-seeking behavior, perceived barriers, and anxiety among adult cancer patients during the COVID-19 pandemic, offering insights into this unique healthcare challenge.

## Methods

### Study design

A cross-sectional study was utilized to explore the health-seeking behavior of adult cancer patients during the COVID-19 pandemic. The research was conducted at two healthcare institutions: the National Institute of Cancer Research and Hospital (NICRH) and Ahsania Mission Cancer and General Hospital, both situated in Dhaka, Bangladesh. The data collection for this study was carried out from August 2020 to December 2020. The study participants encompassed adult cancer patients. The selection of respondents was representative of a broad spectrum of socio-demographic characteristics.

## Selection Criteria

Inclusion Criteria

- Individuals aged 18 years and above, regardless of gender.
- Patients who had been diagnosed with cancer before the onset of the pandemic.

Exclusion Criteria

- Patients who were severely ill
- Respondents who declined to participate in the study.

## Sample Size Estimation

The sample size was estimated using the following formula: n = z²pq/d²

Where,

n = desired sample size

z = confidence interval (1.96 for 95% confidence interval) p = proportion of target population

q = (1 -p)

d = Absolute precision, 5% = (0.05)

Based on the calculation, the desired sample size was n = (1.96) ² x 0.5 x 0.5 / (0.05) ² = 384. To account for non-respondents, the sample size was increased by 10% to 422. Eventually, 210 eligible respondents were interviewed during the defined data collection period.

### Sampling Technique

A purposive sampling technique was utilized for participant selection.

### Data Collection

Data collection was carried out using a pretested semi-structured questionnaire. The questionnaire included questions on socio-demographic characteristics and questions about health-seeking behaviour. We categorized health-seeking behaviour as contact with a health care provider (HCP) since the pandemic, cancelled appointments since the pandemic and Actions taken after cancelling appointments. The questionnaire included questions on socio-demographic characteristics among the respondents, with a cut-off value of 10 [7].

### Data Collection Technique

Data were collected through face-to-face interviews with participants. Before data collection, respondents were provided with comprehensive explanations of the study’s purpose, and written informed consent was obtained. Data collected were thoroughly reviewed and verified daily, rectifying any inaccuracies promptly.

### Pre-Testing

Before the primary data collection, the questionnaire was initially translated into Bangla before use. A pilot test was conducted at Delta Medical College and Hospital in Dhaka. Feedback from the pilot-testing phase led to necessary adjustments and refinements to the questionnaire.

### Data Processing

After data collection, a comprehensive process was followed, including checking, cleaning, editing, compiling, coding, and categorising to ensure data accuracy, consistency, and quality.

### Data Management and Analysis

IBM Statistical Package for Social Science (SPSS) version 26 was employed for data analysis. Furthermore, the findings were divided into categories.

Descriptive and inferential statistical methods include Chi-square, Fisher’s exact, independent sample t-test, one-way ANOVA, and binary logistic regression. The Chi-square test assessed the association between the dependent and independent variables, and Fisher’s exact test was utilized for small sample sizes. The independent sample t-test and one-way ANOVA were used to compare the mean scores. Furthermore, binary logistic regression was carried out to identify the predictors. A significance level of p < 0.05 was considered statistically significant.

### Ethical compliance

Before commencing the study, the Institutional Review Board (IRB) of the National Institute of Preventive and Social Medicine (NIPSOM) approved the research protocol.

### Privacy and informed consent

We upheld the rights of our participants, ensuring that study details were explained in the local language for informed written consent. Private interviews were conducted with utmost confidentiality, and participant concerns were addressed as needed. This approach, which respected the autonomy and privacy of our participants, should make our audience feel that the study was conducted with sensitivity and care.

## Results

Table 1 shows 210 participants in the study, with 56.2% being men and 43.8% being women. Regarding educational attainment, the most significant percentage (36.7%) had never attended school, followed by those who had completed higher secondary education (31.4%). 51.0% of participants were employed, compared to 46.2% of housewives. Among the total participants, 170 of them admitted to postponing appointments. 55.9% followed up with actions after cancelling appointments, whereas 44.1% did not. Regarding the frequency of cancelled appointments, there were no discernible variations according to age groups (p=0.416) or gender (p=0.862). Likewise, actions taken after the cancellation did not significantly differ in gender or age (p=0.199 and p=0.199, respectively). Appointment cancellations were more common among participants who used their vehicle (p=0.105).

**Table 1.** Participant Characteristics and Appointment Cancellation Behavior.

| Variables |  | Contact since the pandemic (always) (n=210) |  |  | Cancelled appointments (yes) (n=170) |  |  | Actions taken after cancelling the appointment. (yes) (n=170) |  |  |
| --- | --- | --- | --- | --- | --- | --- | --- | --- | --- | --- |
|  |  | Sometimes | always | P value | Yes | No | P value | No | Yes | P value |
| **Sex** |  |  |  |  |  |  |  |  |  |  |
| Male | 118(56.2) | 100 (47.7) | 18 (8.5) | 0.416 | 95(45.2) | 23(11.0) | 0.862 | 58(34.1) | 37(21.8) | 0.199 |
| Female | 92(43.8) | 82 (39.0) | 10 (4.8) |  | 75(35.7) | 17(8.1) |  | 53(31.2) | 22(12.9) |  |
| **Religion** |  |  |  |  |  |  |  |  |  |  |
| Islam | 1969(93.3) | 11(5.2) | 3(1.4) | 0.407 | 11(5.2) | 3(1.4) | 0.733 | 6(3.5) | 5(2.9) | 0.517 |
| Hindu | 14(6.7) | 171(81.4) | 25(11.9) |  | 159(75.7) | 37(17.6) |  | 105(61.8) | 54(31.8) |  |
| **Education** |  |  |  |  |  |  |  |  |  |  |
| No formal education | 77(36.7) | 70(33.3) | 7(3.3) | 0.469 | 65(31.0) | 12(5.7) | 0.188 | 46(27.1) | 19(11.2) | 0.648 |
| Primary education | 48(22.9) | 39(18.6) | 9(4.3) |  | 42(20.0) | 6(2.9) |  | 25(14.7) | 17(10.0) |  |
| Secondary education | 19(9.0) | 16(7.6) | 3(1.4) |  | 15(7.1) | 4(1.9) |  | 9(5.3) | 6(3.5) |  |
| Higher secondary | 66(31.4) | 57(27.1) | 9(4.3) |  | 48(22.9) | 18(8.6) |  | 31(18.2) | 17(10.0) |  |
| ry<br>educati<br>on |  |  |  |  |  |  |  |  |  |  |
| **Occupation** |  |  |  |  |  |  |  |  |  |  |
| Unempl<br>oyed/H<br>ousewif<br>e | 97(46.2) | 86(41.0) | 11(5.2) | 0.815 | 77(36.7) | 20(9.5) | 0.888 | 53(31.2) | 24(14<br>.1) | 0.55<br>9 |
| Employ<br>ed | 107(51.0) | 91(43.3) | 16(7.6) |  | 88(41.9) | 19(9.0) |  | 54(31.8) | 34(20<br>.0) |  |
| Student | 6(2.9) | 5(2.4) | 1(0.5) |  | 5(2.4) | 1(0.5) |  | 4(2.4) | 1(0.6) |  |
| **Marital**<br>**status** |  |  |  |  |  |  |  |  |  |  |
| Unmarr<br>ied | 13(6.2) | 10(4.8) | 3(1.4) | 0.389 | 11(5.2) | 2(1.0) | 1.00 | 6(3.5) | 5(2.9) | 0.51<br>7 |
| Married | 197(93.8) | 172(81.9) | 25(11.9) |  | 159(75.7) | 38(18.1<br>) |  | 105(61.8) | 54(31<br>.8) |  |
| **Family**<br>**type** |  |  |  |  |  |  |  |  |  |  |
| Single | 82(39.0) | 73(34.8) | 9(4.3) | 0.534 | 61(29.0) | 21(10.0<br>) | 0.071 | 43(25.3) | 18(10<br>.6) | 0.31<br>7 |
| Joint<br>family | 128(61.0) | 109(51.9) | 19(9.0) |  | 109(51.9) | 19(9.0) |  | 68(40.0) | 41(24<br>.1) |  |
| **Has**<br>**person**<br>**al**<br>**transpo**<br>**rt** |  |  |  |  |  |  |  |  |  |  |
| Yes | 36(17.1) | 28(13.3) | 8(3.8) | 0.105 | 32(15.2) | 4(1.9) | 0.245 | 15(8.8) | 17(10<br>.0) | **0.02**<br>**2** |
| No | 174(82.9) | 154(73.3) | 20(9.5) |  | 138(65.7) | 36(17.1<br>) |  | 96(56.5) | 42(24<br>.7) |  |

GAD score has been demonstrated where the majority (41.4%) of participants suffered from minimal anxiety (reference score 0-4) and 17.6% of participants suffered from moderate anxiety (reference score 10-14).

Participants with personal transportation had a mean GAD score higher than those without, and this difference was statistically significant. However, the table offers no details on the relationship between SD and GAD scores. The participants showed a range of anxiety levels, with a mean age of 42±15 years. Disparities in education showed a significant correlation (p=0.022). Anxiety levels were found to be correlated with higher education levels, indicating a wider range of variability among those with higher secondary education. Anxiety levels and employment status did not significantly correlate in the workplace (>0.05). The mean GAD score was higher in students than unemployed/housewives and employed participants. However, the difference was not statistically significant. Participants with personal transport had a higher mean score of GAD (8.69±6.6) than those who did not have personal transport (7.81±7.3).

**Table 2:**
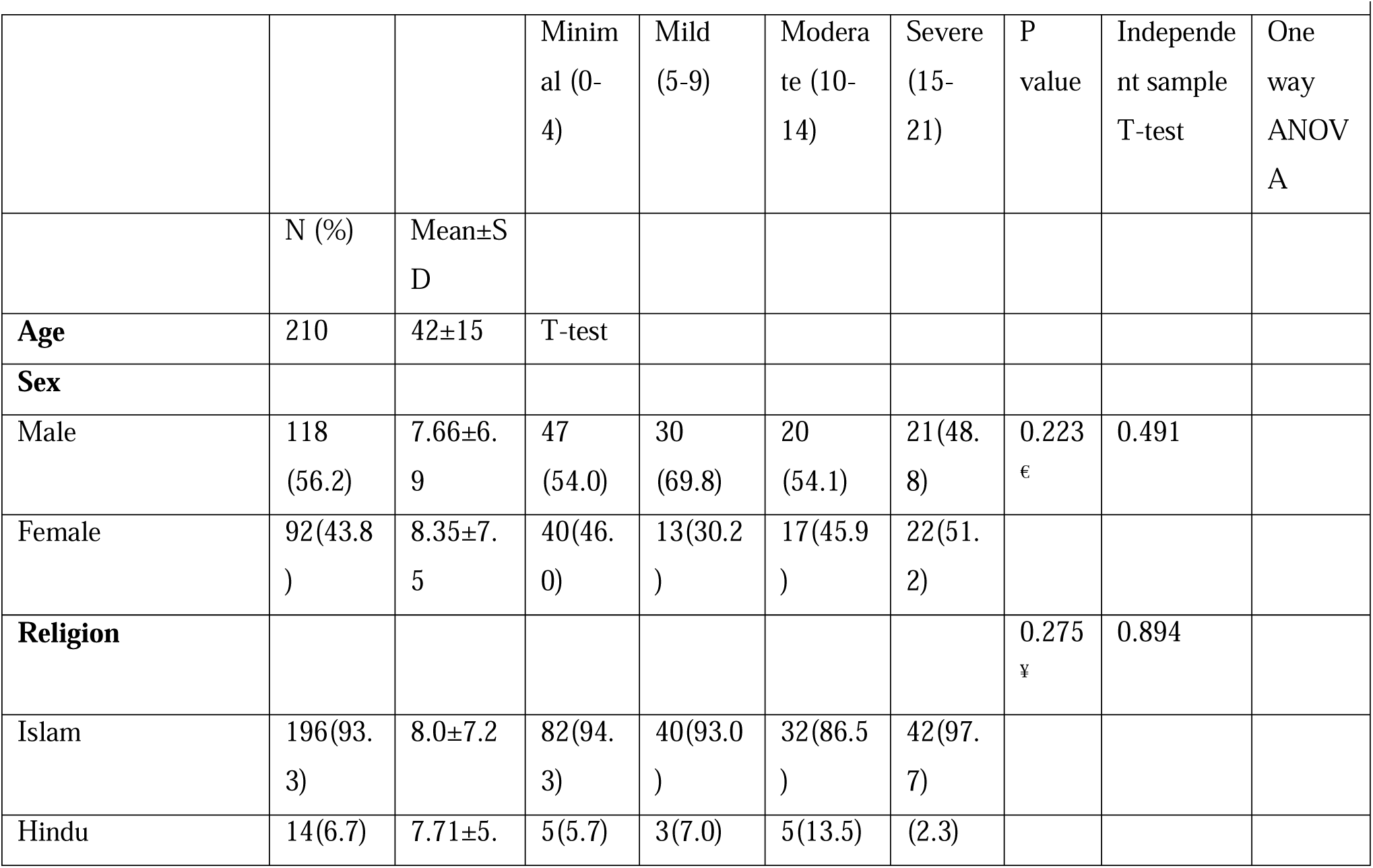
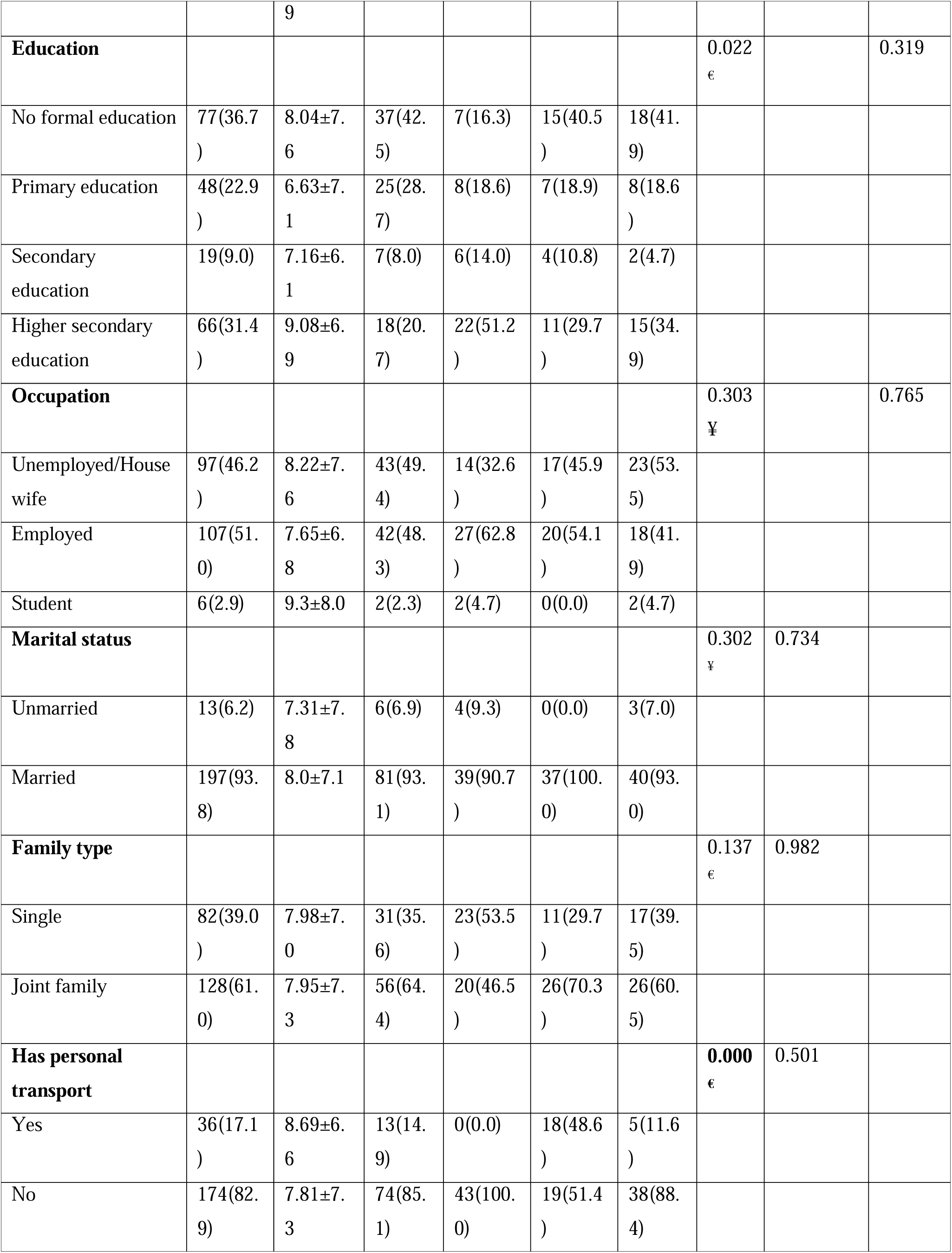
Distribution and Relation between SD and GAD.

Before the outbreak, participants’ interactions with HCP varied in intensity. Remarkably, 64.8% reported regular contact, compared to 35.2% who reported irregular contact. On the other hand, since the outbreak, a smaller subset (13.3%) continued regular contact with HCP after the outbreak, whereas a significant majority (86.7%) reported only sporadic contact. Before the outbreak, participants’ interactions with HCP varied in intensity. Remarkably, 64.8% reported regular contact, compared to 35.2% who reported irregular contact. Since the Outbreak, a smaller subset (13.3%) continued regular contact with HCP after the outbreak, whereas a significant majority (86.7%) reported only sporadic contact.

**Table 3:**
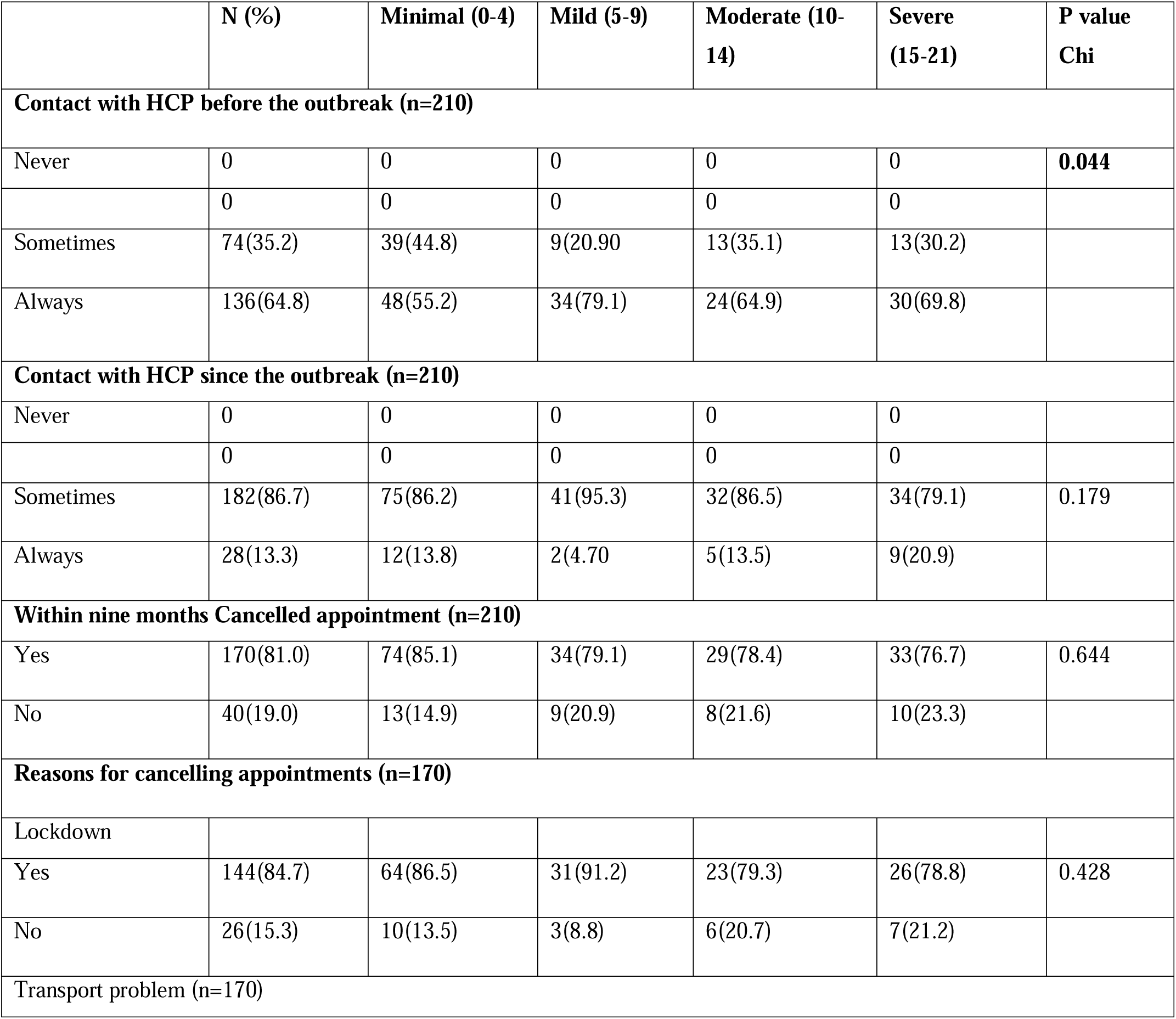
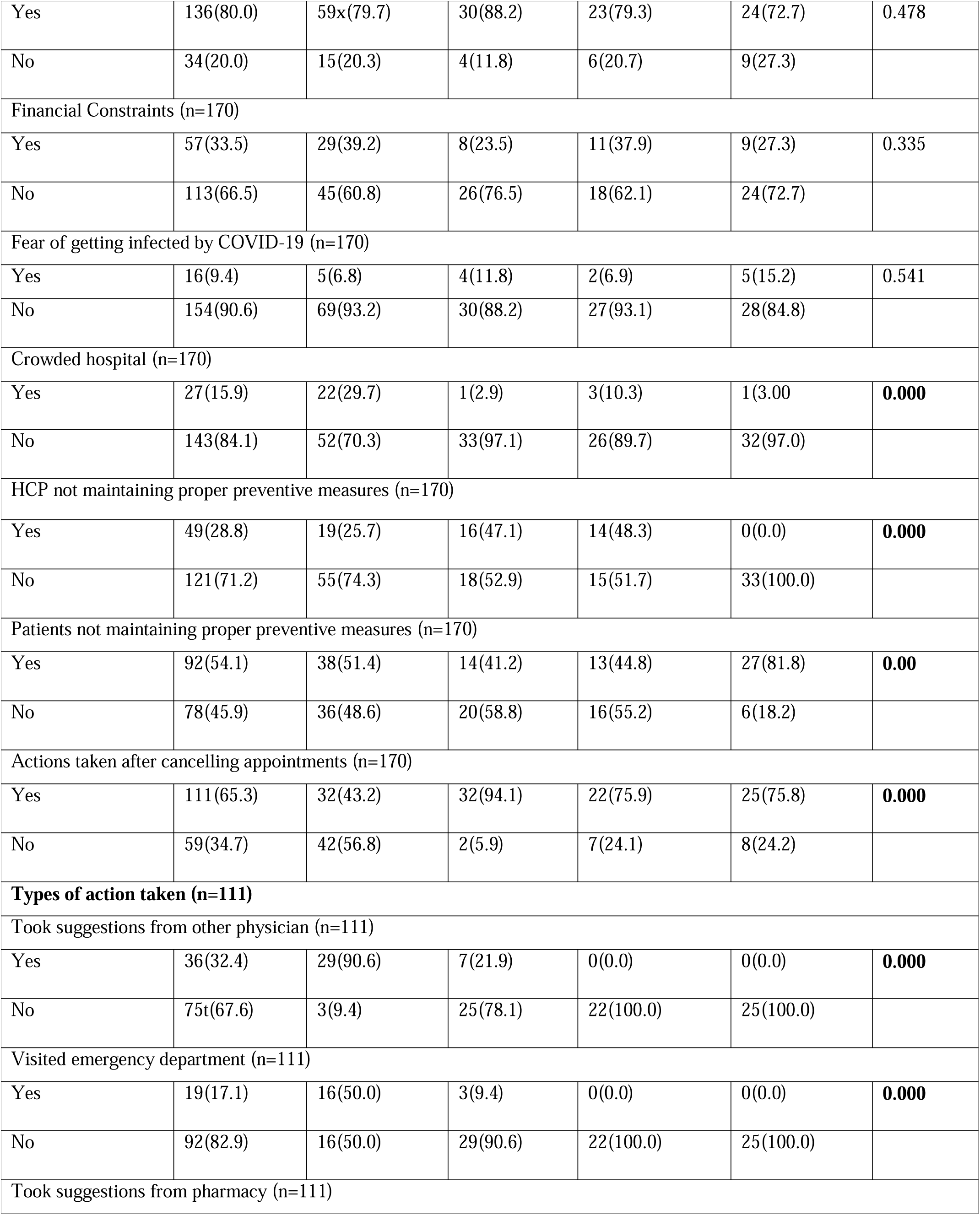
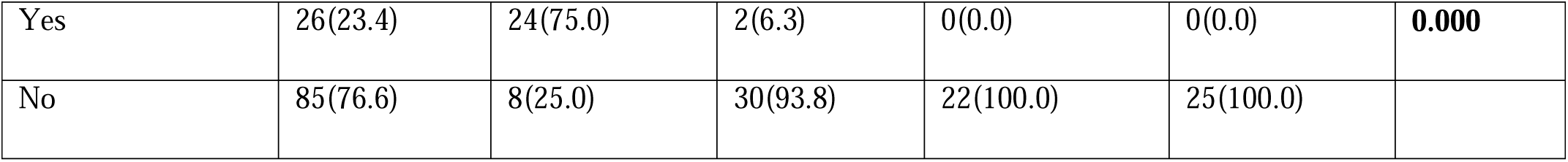
Distribution and Relation between HCP and GAD.

The probability of appointment cancellations was not significantly impacted by age (OR=1.0, 95% CI=1.0-1.1, p=0.163). In contrast to men, women had a greater odds ratio of 1.6 (95% CI=0.1-18.1, p=0.703) for appointment cancellations when gender was taken into account. Compared to those without formal education, those with formal education had 3.2 times higher odds (95% CI=1.0-10.4, p=0.048) of cancelling appointments. Cancellations were not significantly influenced by employment status. Compared to single people, married people showed a lower probability (OR=0.3, 95% CI=0.03-1.9, p=0.182) of calling in sick days. Joint family members also showed a marginally higher probability (OR=1.7, 95% CI=0.6-4.8, p=0.288). Compared to those who had personal transport, participants without it had lower odds (OR=0.5, 95% CI=0.2-1.4, p=0.194) of appointment cancellations.

There is a significant correlation between higher anxiety levels and a higher chance of appointment cancellations. Comparing participants with mild anxiety to those with severe anxiety, there was a significant odds ratio of 3.2 (95% CI=1.4-7.2, p=0.005). When compared to males, females showed a significantly lower likelihood (OR=0.1, 95% CI=0.02-1.0, p=0.045) of acting following cancellations. There was a lower likelihood of action following cancellation among those with formal education. Interestingly, those without access to personal transport had noticeably higher odds of acting later on (OR=4.2, 95% CI=1.7-10.3, p=0.002). An odds ratio of 3.2 (95% CI=1.4-7.2, p=0.005) showed that participants with severe anxiety were more likely to act following cancellations than those with mild anxiety.

**Table 4:**
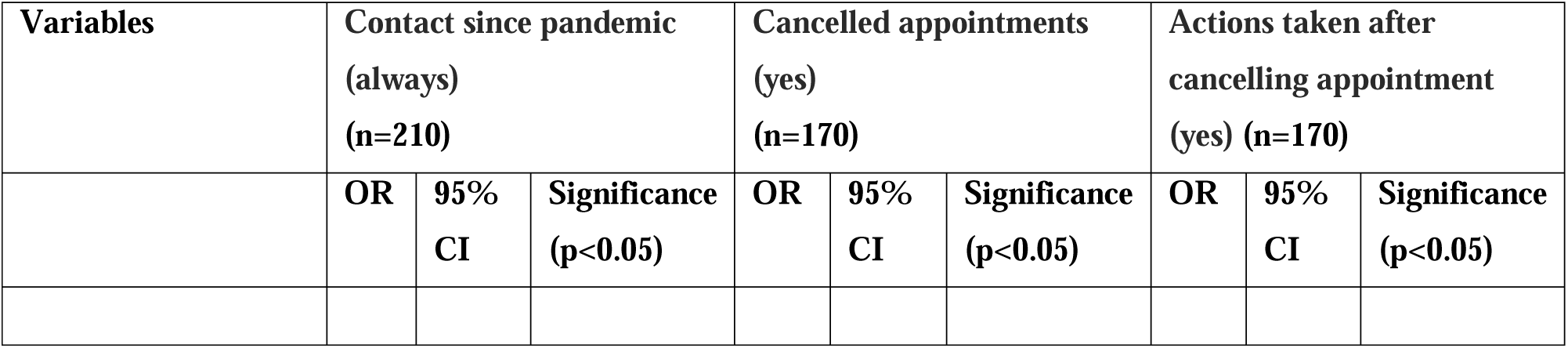
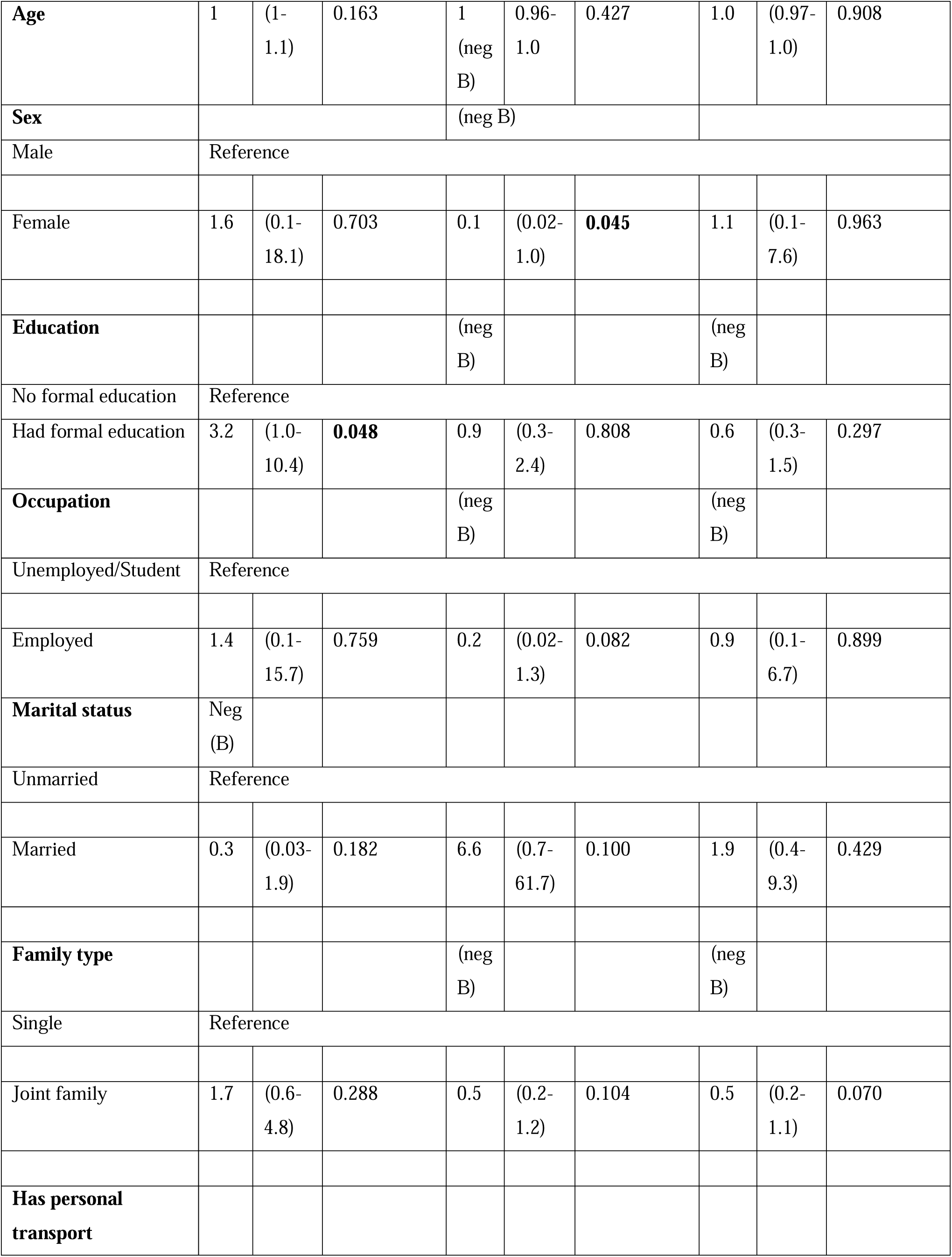
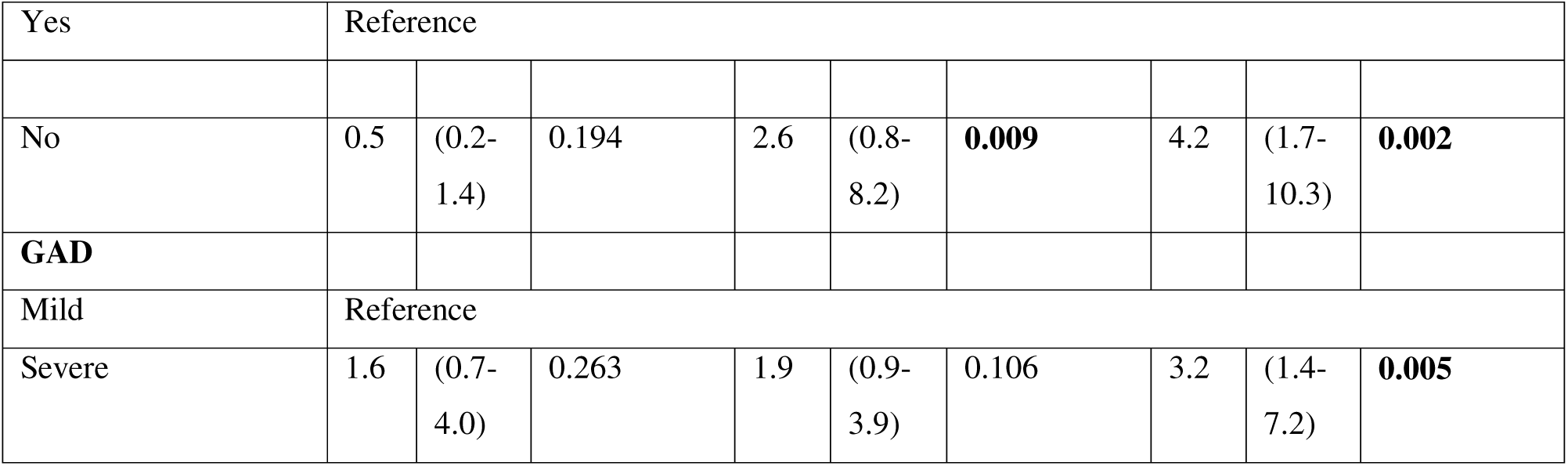
Binary logistic regression between dependent and independent variable.

## Discussion

In this study, we conducted a descriptive cross-sectional analysis among 210 adult cancer patients to assess the relationship between HSB and GAD. The mean age ± SD of the respondents in years was 42 ± 15, and our sample respondents were predominantly male (56.20%). Education levels among our respondents varied, with 36.7% having no formal education while only 31.4% holding a higher secondary education. Comparing our study to similar research, we note some differences. For instance, Tashkandi et al. (2020) found a higher proportion of female participants (61%) with a mean age of 50 ± 15.1 years [8], while Younger et al.’s UK study in 2020 had a median age of 58 years and a higher proportion of females (55%), with the majority being well-educated (73%) [9]. These variations could stem from different sample sizes and socio-demographic factors.

A significant number of the participants in our study showed a low level of anxiety, which aligns with the findings documented in previous studies [10–14]. Usually, strong resilience and effective coping mechanisms are commonly observed among the vulnerable population. Their relatively low level of anxiety during the pandemic may be because they are accustomed to coping with their health issue, which is in and of itself a challenging condition. However, some studies have found higher anxiety levels among cancer patients [15–18]. These differences in the anxiety levels may be attributed to the difference in personal experience of risk perception and psychological flexibility, which leads to different responses to the outside stressors like the pandemic.

Our study observed a significant shift in health seeking behavior while it was focusing on contacting the healthcare providers. A significant number of the respondents with varying levels of anxiety reported a decline in their interactions, with many individuals cancelling their appointments with the HCPs since the pandemic compared to their previous normal routine engagement. The observed decline in interpersonal interactions aligns with patterns identified in previous research, underscoring the significant influence of public health crises on the utilization of healthcare facilities. It is worth noting that this drop was observed among respondents with various levels of anxiety, which is a challenge to the notion that anxiety levels have a direct influence on healthcare-seeking behavior. Another study findings also suggest that cancellation of appointments was associated with low level of anxiety [19]. However, during normal circumstances, individuals with anxiety disorders are more likely to utilize the healthcare services across multiple levels of care [20], and cancelling appointments were associated with high anxiety level [21–22]. However, the decrease in health seeking behaviour seen is consistent with the findings of other studies conducted during COVID-19 pandemic and SARS epidemic [2, 23–25], highlighting the impact of health emergencies on health care seeking behaviour. Surprisingly, the major reasons for cancelling appointments were associated with lockdown and lack of available transportation options, reflecting common challenges in accessing healthcare services, which is in line with the findings of prior studies which have highlighted the impact of mobility restrictions on the individual’s health seeking behaviour during the time of crisis period [26–29]. While several studies highlighted the risk of getting infected by COVID-19 infection, financial constrains or crowded environment in the hospitals as key determents of health seeking behaviour during the pandemic [30–32], our findings suggested that, irrespective of anxiety level, the respondents did not consider these factors as the primary determinants to influence their decision-making process.

Furthermore, our research has shown a statistically significant finding regarding the vital role of HCPs in the lives of the respondents. It was observed that most respondents did not see the potential risk that may arise due to the HCPs’ inability to comply with appropriate preventive measures. This observation suggests a substantial reliance on healthcare providers for their general state of health. Cancer patients frequently place their trust in healthcare professionals (HCPs) because of the HCPs’ specialized knowledge in navigating the complicated nature of the illness and offering psychological assistance. Trust is cultivated by promoting transparent communication, enabling patients to make well-informed choices regarding their medical care, and establishing a nurturing atmosphere that aids patients in managing the difficulties associated with their ailment [33]. It also indicates that individuals diagnosed with cancer tend to prioritize their immediate personal dangers over the hypothetical risk of contracting COVID-19. This highlights the significant faith and dependence put on healthcare professionals’ competence and guidance.

Despite the variation in anxiety levels among respondents, our analysis did not find any significant differences between anxiety levels and the occurrence of appointment cancellations. Potential rationales could encompass extrinsic elements such as challenges related to transportation, or unanticipated events that overshadow the impact of anxiety on the act of canceling appointments. The influence of external circumstances on canceling behaviour may outweigh the impact of worry in this context [34–39].

This result presents a contrast with another study that was focused on pregnant women, where a significantly association was found between health anxiety and the act of appointment cancellations [21]. However, it was observed that GAD showed a statistically significant influence on the subsequent activities undertaken following cancellations. Specifically, those with lower level of anxiety exhibited a higher tendency to actively seek consultation from various healthcare professionals or visiting emergency departments, which aligns with other study findings [40]. Individuals who exhibit reduced levels of anxiety frequently demonstrate an enhanced perception of control and self-assurance in effectively addressing their health-related worries [41–42]. The possession of emotional stability empowers individuals to approach healthcare encounters with a proactive perspective, wherein they actively seek assistance and help from healthcare providers as a strategy for preventive care or prompt intervention [43]. The decrease in their anxiety levels may enable individuals to view the healthcare system as a valuable tool for addressing health-related inquiries and concerns, rather than perceiving it as a cause of more stress or unease. This proactive behavior reflects their efforts to effectively address their health issues in the face of managing their health concerns throughout challenging situations. Moreover, the findings of our research indicate a robust association between academic attainment and the probability of establishing and sustaining a consistent and engaged connection with healthcare practitioners. On the other hand, the research findings indicate that there is a greater probability of appointment cancellations among male participants and individuals who do not have personal transportation during the COVID-19 pandemic. These findings highlight the influence of demographic and logistical factors on healthcare accessibility and the ability to adhere to appointments.

The observed rise in appointment cancellations among male participants may be attributed to various socio-cultural and behavioural factors. Historically, there has been a propensity for males to display hesitance when it comes to seeking medical aid or participating in routine healthcare appointments. This can be attributed to societal norms around masculinity, which often emphasise self-reliance and downplay the significance of health issues. In addition, the existing societal expectations around gender frequently inhibit males from openly expressing their vulnerability or actively seeking prompt medical assistance, which may potentially contribute to a heightened frequency of appointment cancellations [44].

Likewise, the lack of individual conveyance might be a substantial obstacle to attaining healthcare services, particularly in a widespread health emergency like the COVID-19 pandemic. Individuals needing more personal transportation may need help accessing healthcare services, particularly in regions with inadequate public transit or during periods of restricted mobility caused by lockdowns or transportation constraints. The absence of adequate transportation infrastructure might hinder individuals’ capacity to attend scheduled appointments, resulting in an increased probability of cancellations, particularly among individuals dwelling in geographically isolated or underserved regions.

These observations underscore the significance of creating tailored interventions and support strategies that effectively address diverse patient populations’ distinct needs and concerns. Ensuring continuous access to essential healthcare services is paramount, particularly during challenging circumstances.

## Limitations

The present investigation was subject to many limitations. The findings of this study may need more generalizability due to the small sample size employed. Utilising a cross-sectional design in this study may impede the identification of causal linkages and temporal dynamics. The utilisation of self-reported data to evaluate levels of anxiety and healthcare-seeking behaviour presents the potential for response bias and social desirability bias.

## Conclusion

Psychological resilience, environmental constraints, and demographics affect healthcare access and use and complicate patient decision-making. External constraints like mobility restrictions and lockdowns underscore the need for specific interventions and support for high anxiety and diverse demographics. More research is needed to understand the complicated link between anxiety and healthcare-seeking. Policy initiatives should be based on patient decision-making mechanisms to help people navigate complex healthcare environments, increasing health outcomes and equity.

## Data Availability

All data produced in the present study are available upon reasonable request to the authors

## Acknowledgements

Not applicable.

## References

1. UK Coronavirus Cancer Monitoring Project team. The UK Coronavirus Cancer Monitoring Project: protecting patients with cancer in the era of COVID-19. The Lancet. Oncology. 2020 May;21(5):622.

2. Shinan-Altman S, Levkovich I. COVID-19 precautionary behaviour: The Israeli case in the initial stage of the outbreak. BMC public health. 2020 Dec; 20:1–7.

3. Khan S, Jan H, Faisal S, Khan A, Ilyas R, Bibi K, Rehman S. Association of COVID-19 and Cancer: Risk Statistics and Management of COVID-19 in Cancer Patients. International Cardiovascular Forum Journal 2020; 20:5–9.

4. World Health Organization (2020). Naming the coronavirus disease (COVID-19) and the virus that causes it. [online] www.who.int. Available at:

5. https://www.who.int/emergencies/diseases/novel-coronavirus-2019/technical-guidance/naming-the-coronavirus-disease-(covid-2019)-and-the-virus-that-causes-it.

5. Hussain SM. Comprehensive update on cancer scenario of Bangladesh. South Asian J Cancer. 2013 Oct;2(4):279–84.

6. Rath H, Shah S, Sharma G, Mishra E. Exploring determinants of care-seeking behaviour of oral cancer patients in India: A qualitative content analysis. Cancer Epidemiol. 2018 Apr;53:141–148.

7. Spitzer RL, Kroenke K, Williams JBW, Löwe B. A Brief Measure for Assessing Generalized Anxiety Disorder: The GAD-7. Arch Intern Med. 2006;166(10):1092–1097.

8. Tashkandi E, BaAbdullah M, Zeeneldin A, AlAbdulwahab A, Elemam O, Elsamany S, Alfayez M, Dabash Y, Khayat E, Hassanin F, Abdulhameed R. Optimizing the communication with cancer patients during the COVID-19 pandemic: patient perspectives. Patient preference and adherence. 2020 Jul 20:1205–12.

9. Younger E, Smrke A, Lidington E, Farag S, Ingley K, Chopra N, Maleddu A, Augustin Y, Merry E, Wilson R, Benson C. Health-related quality of life and experiences of sarcoma patients during the COVID-19 pandemic. Cancers. 2020 Aug 14;12(8):2288.

10. Anindyajati G, Wiguna T, Murtani BJ, Christian H, Wigantara NA, Putra AA, Hanafi E, Minayati K, Ismail RI, Kaligis F, Savitri AI. Anxiety and its associated factors during the initial phase of the COVID-19 pandemic in Indonesia. Frontiers in psychiatry. 2021 Mar 10;12:634585.

11. van’t Spijker A, Trijsburg RW, Duivenvoorden HJ. Psychological sequelae of cancer diagnosis: a meta-analytical review of 58 studies after 1980. Psychosomatic medicine. 1997 May 1;59(3):280–93.

12. Tsaras K, Papathanasiou IV, Mitsi D, Veneti A, Kelesi M, Zyga S, Fradelos EC. Assessment of Depression and Anxiety in Breast Cancer Patients: Prevalence and Associated Factors. Asian Pac J Cancer Prev. 2018 Jun 25;19(6):1661–1669.

13. Abete-Fornara G, Mameli F, Ruggiero F, Meessen J, Blanda A, Ampollini A, Locatelli M, Salmaggi A, Di Cristofori A, Mauri I, Caroli M. Brain tumours in the time of COVID-19: An online survey on patients’ disease experience in one Italian region. Frontiers in Oncology. 2023 Jan 26;13:1002895.

14. Karataş T, Ayaz-Alkaya S, Özdemir N. Fear, Anxiety, and Coping Self-efficacy of Individuals With Cancer During COVID-19 and Predictive Risk Factors: A Descriptive and Correlational Study. Semin Oncol Nurs. 2023 Aug;39(4):151420.

15. Ayubi E, Bashirian S, Khazaei S. Depression and anxiety among patients with cancer during COVID-19 pandemic: a systematic review and meta-analysis. Journal of Gastrointestinal Cancer. 2021 Jun;52(2):499–507.

16. Alagizy HA, Soltan MR, Soliman SS, Hegazy NN, Gohar SF. Anxiety, depression and perceived stress among breast cancer patients: single institute experience. Middle East Current Psychiatry. 2020 Dec;27:1–0.

17. Hinz A, Herzberg PY, Lordick F, Weis J, Faller H, Brähler E, Härter M, Wegscheider K, Geue K, Mehnert A. Age and gender differences in anxiety and depression in cancer patients compared with the general population. European journal of cancer care. 2019 Sep;28(5):e13129.

18. Miaskowski C, Paul SM, Snowberg K, Abbott M, Borno H, Chang S, Chen LM, Cohen B, Hammer MJ, Kenfield SA, Kober KM. Stress and symptom burden in oncology patients during the COVID-19 pandemic. Journal of pain and symptom management. 2020 Nov 1;60(5):e25–34.

19. Steel JL, Amin A, Peyser T, Olejniczak D, Antoni M, Carney M, Tillman E, Hecht CL, Pandya N, Miceli J, Reyes V. The benefits and consequences of the COVID 19 pandemic for patients diagnosed with cancer and their family caregivers. Psycho Oncology. 2022 Jun;31(6):1003–12.

20. Horenstein A, Heimberg RG. Anxiety disorders and healthcare utilization: A systematic review. Clinical psychology review. 2020 Nov 1;81:101894.

21. Shayganfard M, Mahdavi F, Haghighi M, Sadeghi Bahmani D, Brand S. Health Anxiety Predicts Postponing or Cancelling Routine Medical Health Care Appointments among Women in Perinatal Stage during the Covid-19 Lockdown. Int J Environ Res Public Health. 2020 Nov 9;17(21):8272.

22. Voisin MR, Oliver K, Farrimond S, Chee T, Arzbaecher J, Kruchko C, Maher ME, Tse C, Cashman R, Daniels M, Mungoshi C. Brain tumors and COVID-19: the patient and caregiver experience. Neuro-oncology advances. 2020 Jan;2(1):vdaa104.

23. Chang HJ, Huang N, Lee CH, Hsu YJ, Hsieh CJ, Chou YJ. The impact of the SARS epidemic on the utilization of medical services: SARS and the fear of SARS. American journal of public health. 2004 Apr;94(4):562–4.

24. Riera R, Bagattini ÂM, Pacheco RL, Pachito DV, Roitberg F, Ilbawi A. Delays and disruptions in cancer health care due to COVID-19 pandemic: systematic review. JCO Global Oncology. 2021 Feb;7(1):311–23.

25. Papautsky EL, Hamlish T. Patient-reported treatment delays in breast cancer care during the COVID-19 pandemic. Breast cancer research and treatment. 2020 Nov;184:249–54.

26. Aklilu TM, Abebe W, Worku A, Tadele H, Haile T, Shimelis D, Mekonen D, Amogne W, Moges A, Habtamu A, Argaw R. The impact of COVID-19 on care seeking behavior of patients at tertiary care follow-up clinics: a cross-sectional telephone survey. Addis Ababa, Ethiopia. medRxiv. 2020 Nov 29:2020–11.

27. Hossain MR, Parray AA, Sultana R, Aktar B, Rashid SF. Exploring healthcare-seeking behavior of most vulnerable groups amid the COVID-19 pandemic in the humanitarian context in Cox’s Bazar, Bangladesh: Findings from an exploratory qualitative study. PLOS Glob Public Health. 2023 Mar 20;3(3):e0000382.

28. Tandon T, Dubey AK, Dubey S, Arora E, Hasan MN. Effects of COVID-19 pandemic lockdown on medical advice seeking and medication practices of home-bound non-COVID patients. J Educ Health Promot. 2021 Jan 28;10:28.

29. Coetzee BJ, Kagee A. Structural barriers to adhering to health behaviours in the context of the COVID-19 crisis: considerations for low-and middle-income countries. Global Public Health. 2020 Aug 2;15(8):1093–102.

30. Chang WH. The influences of the COVID-19 pandemic on medical service behaviors. Taiwanese Journal of Obstetrics and Gynecology. 2020 Nov 1;59(6):821–7.

31. Kirby A, Drummond FJ, Lawlor A, Murphy A. Counting the social, psychological, and economic costs of COVID-19 for cancer patients. Support Care Cancer. 2022 Nov;30(11):8705–8731.

32. Hsieh YP, Yen CF, Wu CF, Wang PW. Nonattendance at scheduled appointments in outpatient clinics due to COVID-19 and related factors in Taiwan: a health belief model approach. International Journal of Environmental Research and Public Health. 2021 Apr 22;18(9):4445.

33. Hillen MA, De Haes HC, Smets EM. Cancer patients’ trust in their physician—a review. Psycho oncology. 2011 Mar;20(3):227–41.

34. Bambra, C., Gibson, M., Sowden, A., Wright, K., Whitehead, M., & Petticrew, M. Tackling the wider social determinants of health and health inequalities: evidence from systematic reviews. Journal of Epidemiology and Community Health, 2010, 64(4), 284–291.

35. Roberge, D., Beaulieu, M. C., & Haddad, S. Long-term effects of a quasi-experimental study to increase patients’ attendance to primary care appointments. Family Practice, 2013, 30(5), 570–578.

36. Stewart, M. A. Effective physician-patient communication and health outcomes: a review. CMAJ: Canadian Medical Association Journal, 1995, 152(9), 1423–1433.

37. Jerant, A., Kravitz, R. L., Azari, R., White, L. F., & Wood, R. Provider characteristics strongly influence patient adherence to chronic disease management. Journal of the American Board of Family Medicine, 2006, 19(5), 494–502.

38. Indini A, Pinotti G, Artioli F, Aschele C, Bernardi D, Butera A, Defraia E, Fasola G, Gamucci T, Giordano M, Iaria A, Leo S, Ribecco AS, Rossetti R, Savastano C, Schena M, Silva RR, Grossi F, Blasi L. Management of patients with cancer during the COVID-19 pandemic: The Italian perspective on the second wave. Eur J Cancer. 2021 May;148:112–116.

39. Indini A, Pinotti G, Artioli F, Aschele C, Bernardi D, Butera A, Defraia E, Fasola G, Gamucci T, Giordano M, Iaria A, Leo S, Ribecco AS, Rossetti R, Savastano C, Schena M, Silva RR, Grossi F, Blasi L. Management of patients with cancer during the COVID-19 pandemic: The Italian perspective on the second wave. Eur J Cancer. 2021 May;148:112–116.

40. Siette J, Seaman K, Dodds L, Ludlow K, Johnco C, Wuthrich V, Earl JK, Dawes P, Strutt P, Westbrook JI. A national survey on COVID-19 second-wave lockdowns on older adults’ mental wellbeing, health-seeking behaviours and social outcomes across Australia. BMC geriatrics. 2021 Dec;21:1–6.

41. Schroevers MJ, Ranchor AV, Sanderman R. The role of social support and self-esteem in the presence and course of depressive symptoms: a comparison of cancer patients and individuals from the general population. Social science & medicine. 2003 Jul 1;57(2):375–85.

42. Sowislo JF, Orth U. Does low self-esteem predict depression and anxiety? A meta-analysis of longitudinal studies. Psychological bulletin. 2013 Jan;139(1):213.

43. Birks YF, Watt IS. Emotional intelligence and patient-centred care. J R Soc Med. 2007 Aug;100(8):368–74.

44. Novak JR, Peak T, Gast J, Arnell M. Associations Between Masculine Norms and Health-Care Utilization in Highly Religious, Heterosexual Men. Am J Mens Health. 2019 May-Jun;13(3):1557988319856739

